# Longitudinal Trend Monitoring of Multiple Sclerosis Ambulation using Smartphones

**DOI:** 10.1101/2022.02.21.22270745

**Authors:** Andrew P. Creagh, Frank Dondelinger, Florian Lipsmeier, Michael Lindemann, Maarten De Vos

**Affiliations:** Institute of Biomedical Engineering, University of Oxford, UK; Roche Innovation Center, Basel, CH; Department of Electrical Engineering, KU Leuven, BE

**Keywords:** Gait, deep learning, multiple sclerosis, digital biomarkers, smartphones

## Abstract

**Goal:** Smartphone and wearable devices may act as powerful tools to remotely monitor physical function in people with neurodegenerative and autoimmune diseases from out-of-clinic environments. Detection of progression onset or worsening of symptoms is especially important in people living with multiple sclerosis (PwMS) in order to enable optimally adapted therapeutic strategies. MS is a disease whose symptoms typically follow subtle and fluctuating disease courses, patient-to-patient, and over time. Current in-clinic assessments are often too infrequently administered to reflect longitudinal changes in MS impairment that impact daily life. This work, therefore, explores how smartphones can administer daily two-minute walking assessments to monitor PwMS physical function at home.

**Methods:** Remotely collected smartphone inertial sensor data was transformed through state-of-the-art Deep Convolutional Neural Networks, to estimate a participant’s daily ambulatory-related disease severity, longitudinally over a 24-week study.

**Results:** This study demonstrated that smartphone-based ambulatory severity outcomes could accurately estimate MS level of disability, as measured by the EDSS score (*r*^2^: 0.56,*p <*0.001). Furthermore, longitudinal severity outcomes were shown to accurately reflect individual participants’ level of disability over the study duration.

**Conclusion:** Smartphone-based assessments, that can be performed by patients from their home environments, could greatly augment standard in-clinic outcomes for neurodegenerative diseases. The ability to understand the impact of disease on daily-life between clinical visits, through objective digital outcomes, paves the way forward to better measure and identify signs of disease progression that may be occurring out-of-clinic, to monitor how different patients respond to various treatments, and to ultimately enable the development of better, and more personalised care.

## 1 Introduction

Neurodegenerative diseases, such as multiple sclerosis (MS), frequently fluctuate over time, and patient-to-patient, ensuring that it is notoriously difficult to quantify effective therapeutic interventions and disease management techniques. Current in-clinic assessments are often too infrequent to track changes in MS impairment over time. Importantly, it has been shown that earlier identification of changes in PwMS impairment are important to identify and provide better therapeutic strategies [1]. As a result, there exists a great opportunity to augment current clinical examination strategies, to integrate methods that accurately and remotely monitor disease-related changes and deterioration, that may occur at home and between clinician visits.

Although MS follows a highly heterogeneous and subject-specific disease course, the disease profiles can be grouped into four clinical phenotypes which are based on disease progression [2], [3]: the majority of PwMS will initially experience Relapsing–remitting MS (RRMS), a state dominated by sudden acute symptoms developing (a “relapse”) over days before generally plateauing over weeks or months [4], termed “remission”. RRMS generally affects 85% of PwMS and disease activity typically occurs acutely at a sub-clinical level. Secondary-progressive MS (SPMS) can occur in some RRMS patients, where the disease course continues to worsen with or without periods of remission. Half of RRMS patients will go onto develop SPMS [5]–[7]. Those experiencing consistent but worsening symptoms can be thought of as having Primary-progressive MS (PPMS) [4], [5], [7] (roughly 10% of PwMS [6]). Progressive-relapsing MS is more rare (affecting fewer than 5% of PwMS); it occurs from diagnoses as a progressive disease course, with periods of relapse, but without any remission periods.

Digital smartphone-based assessments offer the ability to objectively monitor disability levels in people with multiple sclerosis (PwMS) from out-of-clinic, at home environments [8]–[11]. For instance, smartphone-based monitoring was exemplified in a recent investigation by Bove *et al*. (2015) [12], with this study demonstrating the feasibility of administering daily smartphone-based tasks to PwMS over a one-year period. These technologies can provide new data-driven metrics for clinical decision-making during in-clinic visits [13] and may be more accurate than conventional clinical outcomes, recorded at infrequent visits, to detect subtle, progressive, sub-clinical changes or trends in long-term PwMS disability [12].

Alterations during ambulation (gait) due to MS are a amongst the most common indication of MS impairment [14]–[19]. It has been shown that gait impairment affects quality of life, health status and productivity [20] in persons with MS (PwMS), with the prevalence of these reported impairments between 75% and 90% [21]. PwMS can display postural instability [15], gait variability [16]–[18] and fatigue [19] during various stages of disease progression. The gold-standard assessment of overall disability in MS is the Expanded Disability Status Scale (EDSS) [22], however there are specific functional domain assessments such as the Timed 25-Foot Walk (T25FW), which is part of the Multiple Sclerosis Functional Composite score [23], [24], and the Two-Minute Walk Test (2MWT) which also assesses physical gait function and fatigue in PwMS [25]. In recent years however, there has been a shift towards the adoption of body worn sensors to objectively evaluate ambulatory performance in PwMS, circumventing the need for resource-intensive and expensive gait laboratory equipment, but also opening up the possibility to measure physical function outside of standard clinical settings [13], [17], [18], [26]–[31]

This study builds upon our previous investigations [32]–[34], where we have shown how inertial sensors contained within consumer-based smartphones can be used to characterise gait impairments in PwMS from a remotely administered Two-Minute Walk Test (2MWT). The latter study first introduced how state-of-the-art Deep Convolutional Neural Networks (DCNN) can be applied to remote 2MWT smartphone sensor data to determine a study participants’ status: such as healthy, PwMS with mild, or PwMS with moderate disability. The work presented here aims to evaluate how these DCNN severity predictions from daily 2MWTs can characterise the status of healthy participants versus PwMS with mild, or PwMS with moderate disability over a 24 week period.

## 2 Methods

### 2.1 Data

The FLOODLIGHT (FL) proof-of-concept (PoC) app was trialled in a 24-week, prospective study in PwMS and HCs (NCT02952911) to assess the feasibility of remote patient monitoring using smartphone (and smartwatch) devices [11], [35]. A total of 97 participants (24 HC subjects; 52 mildly disabled, PwMSmild, EDSS [0-3]; 21 moderately disabled PwMSmod, EDSS [3.5-5.5]) contributed data which was recorded from a 2MWT performed out-of-clinic [32]. Subjects were requested to perform a 2MWT daily over a 24-week period, and were clinically assessed at baseline, week 12 and week 24. For further information on the FL dataset and population demographics we direct the reader to [35] and specifically to our previous work [32], [34], which this study expands upon. Table 1 depicts the population demographics for this study.

**Table 1:**
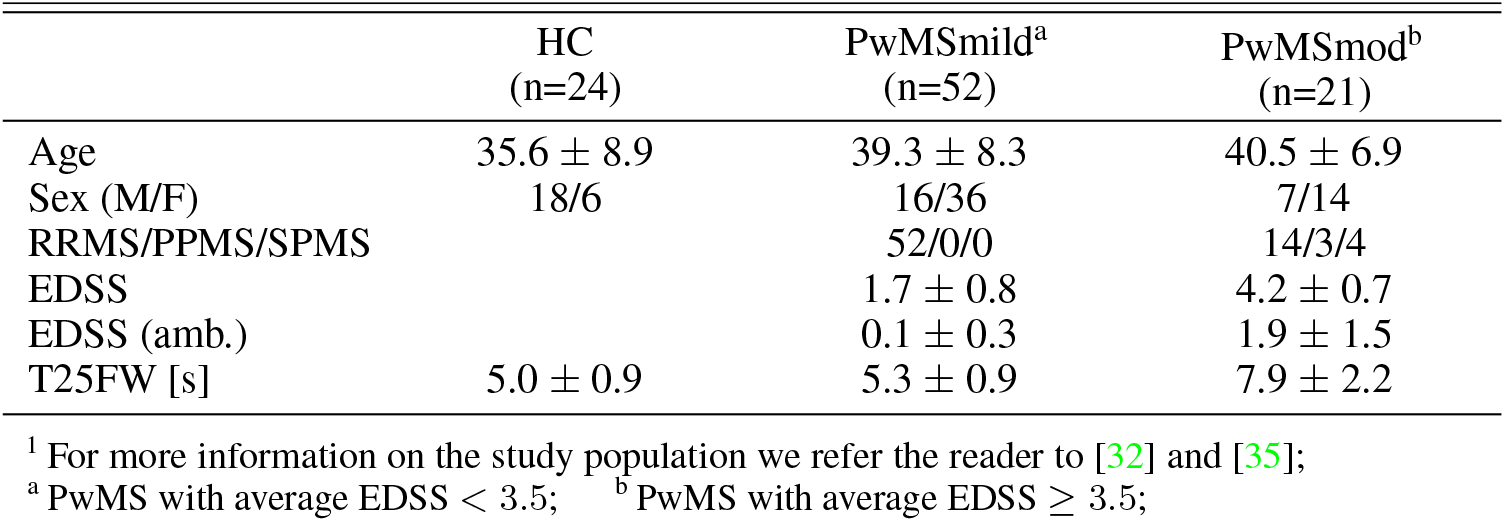
Population Demographics^1^. Clinical scores taken as the average per subject over the entire study, where the mean ± standard deviation across population are reported; RRMS, Relapsing-remitting MS; PPMS, Primary-progressive MS; SPMS, Secondary-progressive MS; EDSS, Expanded Disability Status Scale; T25FW, the Timed 25-Foot Walk; EDSS (amb.) refers to the ambulation sub-score as part of the EDSS; [s], indicates measurement in seconds;

|  | HC<br>(n=24) | PwMSmild<sup>a</sup><br>(n=52) | PwMSmod<sup>b</sup><br>(n=21) |
| --- | --- | --- | --- |
| Age | 35.6 ± 8.9 | 39.3 ± 8.3 | 40.5 ± 6.9 |
| Sex (M/F) | 18/6 | 16/36 | 7/14 |
| RRMS/PPMS/SPMS |  | 52/0/0 | 14/3/4 |
| EDSS | | 1.7 ± 0.8 | 4.2 ± 0.7 |
| EDSS (amb.) | | 0.1 ± 0.3 | 1.9 ± 1.5 |
| T25FW [s] | 5.0 ± 0.9 | 5.3 ± 0.9 | 7.9 ± 2.2 |

<sup>1</sup> For more information on the study population we refer the reader to [32] and [35];
<sup>a</sup> PwMS with average EDSS < 3.5; <sup>b</sup> PwMS with average EDSS ≥ 3.5;

### 2.2 Estimating Ambulatory-related Disease Severity from Smartphone Sensor Data

Smartphone inertial sensor data was recorded while participants performed a daily, at home, two minute walk test (2MWT). The raw sensor data from each 2MWT were then partitioned into multiple vector sequences (epochs), of 2.56 sec (128 samples/epoch) with 50% overlap between adjacent windows. A Deep Convolutional Neural Network (DCNN) was then trained to classify a given epoch as having been performed by a HC, PwMSmild or PwMSmod participant. The DCNN model implemented has previously been introduced in [34], where the network was first pre-trained on the UCI smartphone-based Human Activity Recognition (HAR) dataset, and thereafter fine-tuned on the data in FL for MS severity classification. Briefly, a DCNN applied a series of one-dimensional kernels on the raw sensor epoch **x**_*n*_ with an input (channel 1-4): **X**_*n*_ = (***a***_*x*_, ***a***_*y*_, ***a***_*z*_, ∥***a***∥)^⊤^, where ***a*** are acceleration vectors for the *x*-, *y*- and *z*-components containing samples ***a*** = (*x*_1_, *x*_2_, …, *x*_*T*_) and ∥***a***∥ refers to original orientation invariant signal magnitude. The DCNN consisted of four causal convolutional blocks with batch normalisation (BN) layers (*momentum* = 0.99, *ϵ* = 1*e*^−2^): the 1^st^ block extracted 32 fixed filters with a width of 9 samples, stride length of 1 (9 × 1), with *l*_2_-norm regularisation (*λ* = 1*e*^−3^); the 2^nd^ and 3^rd^ blocks learned 64 filters, with width (3 × 1); the 4^th^ block learned 128 filters with a width of 6 (6 × 1), followed by a final 3-class dense fully connected softmax layer. Max pooling operations were also applied in the 2^nd^ and 4^th^ layers with pool size p=2 and down-scaled by stride factor s=2. The DCNN was trained to minimise a multi-class categorical cross-entropy loss function for *k* ∈ {*hc*}, *mild, mod* to learn the optimal network weights **w**, using an *Adam* optimization algorithm with a learning rate *lr* = 1*e* − 5, as well as *β*_1_ = 0.9 and *β*_2_ = 0.999 which determined the exponential decay rates for the moment estimates of the gradient [36], [37]. The network outputs are interpreted as *ŷ*_*k*_(**x**_*n*_, **w**) = *p*(*y*_*k*_ = *k*|**x**_*n*_). As such, *ŷ*_*k*_ can be thought of as the probability that a given epoch **x**_*n*_ belonged to class *k*. A continuous estimate of severity, the predicted level of MS disability, can then be captured by taking an average of all epoch predictions over a test for a given assessment day, *d* such that:

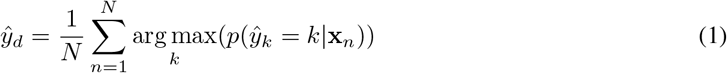

where *N* are the number of windowed epochs for a given test date, *d*, and *k* lies in an ordinal range of [0, 1, …*K*]. Therefore *ŷ*_*d*_ will be continuous such that 0 ≤ *ŷ*_*d*_ ≤ *K* and can conceptualised as a naïve estimate of MS disease severity, mapping a predicted level of disability ranging from healthy to mild to moderate.

Models were trained using a stratified 5-fold cross-validation (CV), with subjects randomly partitioned into one of k=5 folds, as described previously in [32]. One set was denoted the training set (in-sample), which was further split into a smaller set for validation, using roughly 10% of the training subjects. Predictions were evaluated on all available 2MWTS per subject in each of the (out-of-sample) test sets.

### 2.3 Longitudinal Trend Monitoring of Remote Smartphone-Based Outcomes

Longitudinal trends of specific participants were examined as a time-series by considering the severity estimates *ŷ* of repeated 2MWTs over all their available data for the duration of the FL study. While participants were requested to perform a daily Two-Minute Walk Test (2MWT), some test-dates may be missing; it was also observed that various participants had differing adherence rates during the study. There are many strategies to overcome sparsity in clinical data [38], [39]; while some methods aim to impute feature values and raw data, other techniques aim to account for missingness within the construction of a model itself [40], [41]. As the goal of this work was to perform longitudinal analysis of subject’s severity, namely the severity trends over time, missing 2MWT outcomes were imputed by considering *ŷ* as a time-series. Piecewise Linear Interpolation (PLI), was used in this study to impute missing test severity observations on a given date. Other simple and efficient interpolation methods also exist such as linear or previous neighbour interpolation, as well as cubic or spline interpolation [38], [39].

A simple trend estimation can be applied to the time sequence of severity estimates (*ŷ*) across days (*d*) using a *d*− centred linear moving average filter (MAF).

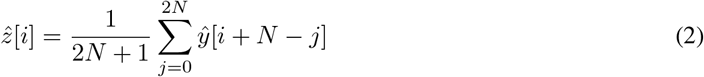

where *ŷ*[·] is the input sequence (severity estimates) and 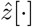 is the output (filtered) sequence (moving severity estimate) for each *d*^*th*^ day; 2*N* + 1 defines the order of the filter, in this case the number of days *d* used in the moving average. A 7-day window was implemented in order to capture the trends in *ŷ*_*d*_ over the study duration.

### 2.4 Statistical Analysis

The association between estimated continuous disease severity and EDSS was tested using (linear) Pearson’s (*r*) and (non-linear) Spearman’s (*ρ*) correlation coefficient. A non-parametric Kruskal-Wallis (KWt) test by ranks assessed the median severity estimate between HC, PwMSmild, and PwMSmod groups. Statistical differences in smartphone severity estimates were also investigated within participants over the duration of the study. For instance, mean differences in severity estimates before and after specific events, such as the reporting of a relapse, were assessed using a *t*-test. In cases where severity estimates had unequal variances before/after an event, as determined by a Brown-Forsythe (BF) test by medians [42], a Welch’s *t*-test correction was applied. Furthermore, differences in median severity estimates before/after each event were also assessed with a non-parametric Mann-Whitney U test.

## 3 Results

### 3.1 Digitally Estimated Severity Outcome

A continuous disease severity outcome was created by averaging all 2MWT predictions (i.e. HC, PwMSmild, PwMSmod) for each participant, calculated from each of the out-of-sample test sets during cross-validation. A disease severity outcome therefore mapped a posterior probability ranging from healthy, to mild, and to moderate for each subject. The distribution of the average severity per subject was displayed in figure 2, and demonstrated the positive relationship between severity outcome and average EDSS, (Pearson’s *r* : 0.75; Spearman’s *ρ* : 0.71; *p <* 0.001, *r*^2^ : 0.56, *p <* 0.001).

**Figure 1:**
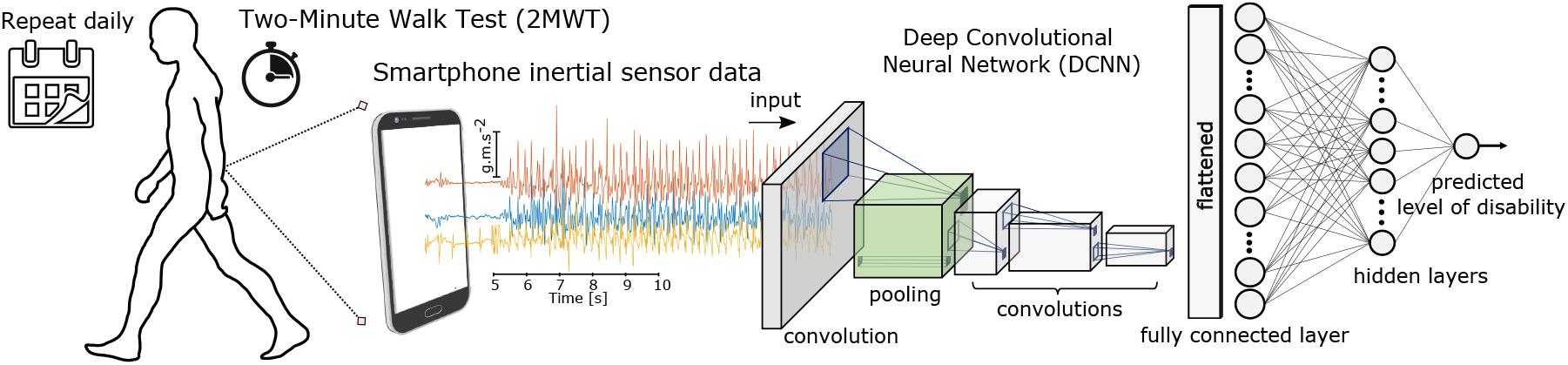
Demonstration of how deep learning algorithms can transform smartphone measurements to predict MS patient severity symptoms between clinical visits. Illustration of Deep Convolutional Neural Network (DCNN) applied to raw smartphone inertial sensor data collected from a remotely executed Two-Minute Walk Test (2MWT), performed daily for 24-weeks.

**Figure 2:**
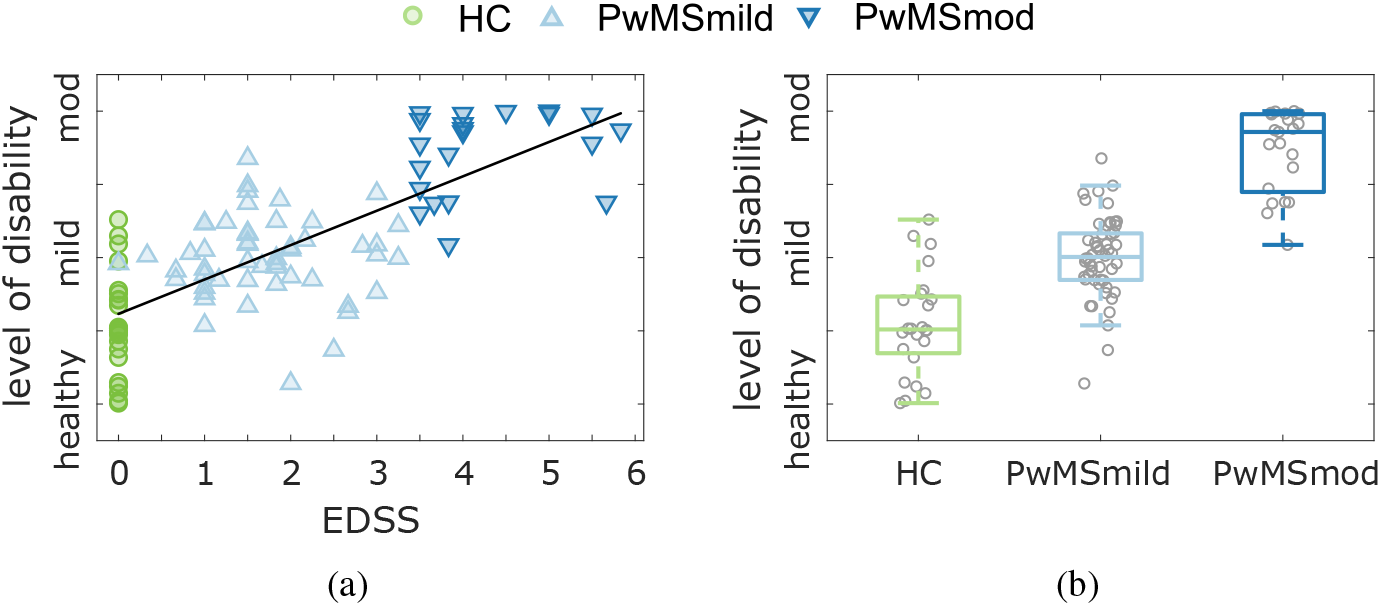
The relationship between the continuous disease severity outcome estimate, EDSS and subject group. Figure **(a)** depicts the scatter plot demonstrating the positive correlation (*r* : 0.75; *ρ* : 0.71; *p <* 0.001) between the average severity outcome and EDSS score per subject; whereas **(b)** represents the distribution of the average severity outcomes per each subject group. A DCNN model was constructed based on the average class predictions (HC, PwMSmild, PwMSmod) per subject over all 2MWTs, creating an estimated continuous prediction probability distribution, ranging from healthy to moderate MS. Each point therefore represents the average estimated severity outcome (probability) for that subject. A black line represents the line of best fit between severity and EDSS (*r*^2^ : 0.56, *p <* 0.001).

### 3.2 Longitudinal Characterisation of Digitally Estimated Severity Outcomes

Disease severity outcomes were evaluated for each 2MWT performed per subject. As a result, longitudinal trends in ambulatory impairment can be monitored by examining daily 2MWT estimates for participants over the duration of the FL study. While the 24-week duration of the study and relatively low level of baseline impairment of the participants meant that we did not observe meaningful progression at the study cohort level, we could still investigate the ability of our methodology to capture participant-specific longitudinal trends. For example, figure 3 examines the longitudinal severity estimate outcome for for various representative correctly classified HC, PwMSmild and PwMSmod participants. Individual 2MWT ambulatory severity estimates are depicted from the 0^*th*^ week until study completion in week 24, where dashed black lines represented site-visits where participants were assessed clinically. Blue lines depicted the 7-day average trend in severity outcomes.

**Figure 3:**
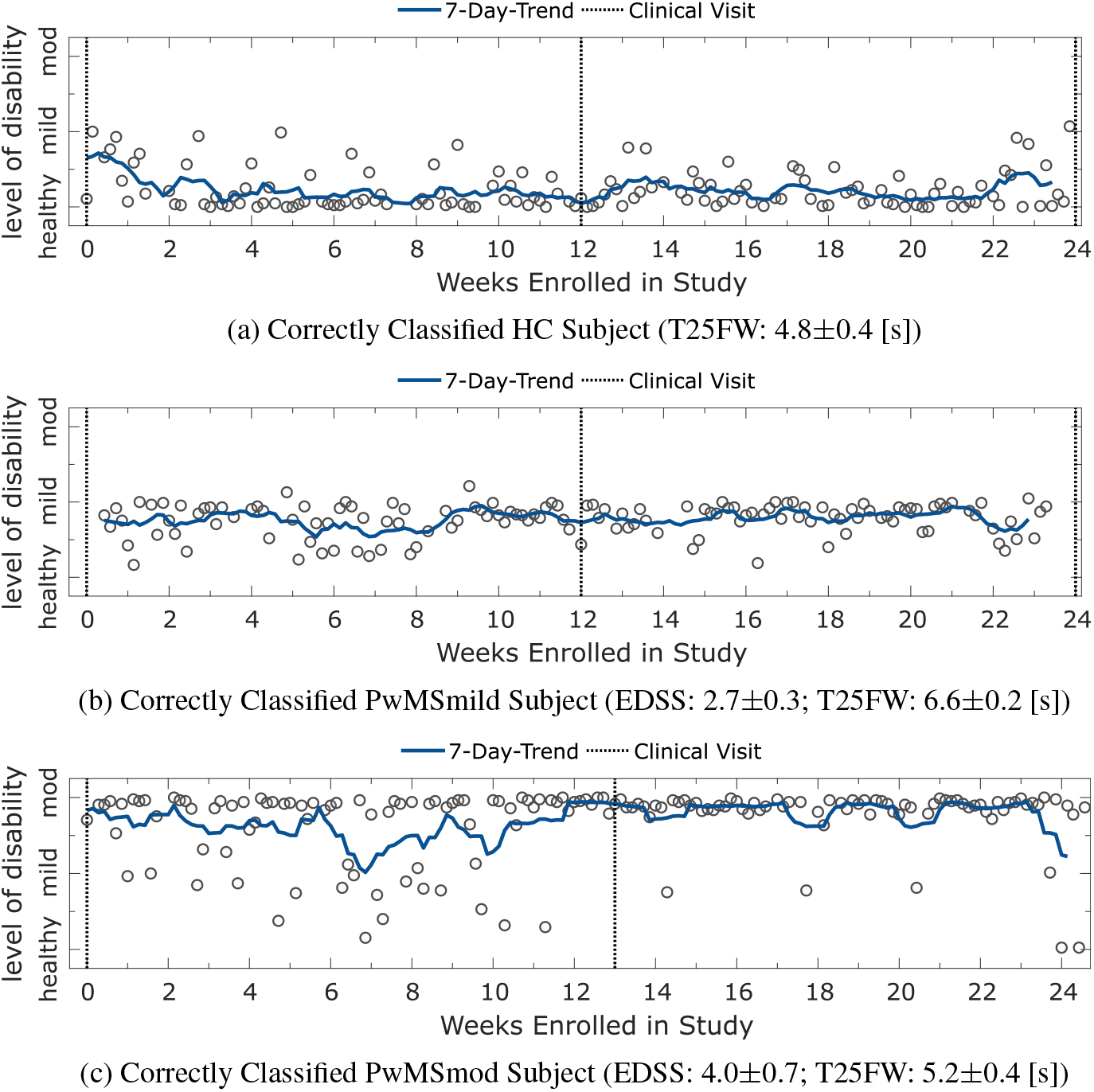
Panel plot illustrating the longitudinal severity estimate outcome for correctly classified HC, PwMSmild and PwMSmod subjects. Depicted are the estimated level of disability for an example **(a)** HC subject; **(b)** a PwMSmild subject; **(c)** a PwMSmod subject during the study. Each circle represents the severity outcome estimate for a 2MWT performed on a given date. Shaded blue lines depict the and 7-day trend, represented by the *d*-point centred moving average across days (*d*). Missing test dates (which are not depicted) were imputed using piecewise linear interpolation. Dashed black lines represent site-visits where the participant was assessed clinically.

Figure 3a first depicted a HC subject. This participant was examined at baseline (week 0; EDSS 0, T25FW: 5 [s]), midway through the study (week 12; EDSS 0^3^; T25FW: 4.5 [s]) and at the study completion (week 24; EDSS: 0; T25FW: N/A^4^ [s]). It was observed that this subject was predicted as healthy with a low severity, consistently across the entire study. Many variations in severity outcomes were smoothed out across the 7-day moving average. Similarly, figure 3b demonstrated a correctly classified, stable, PwMSmild participant over the duration of the study. This participant was also clinically examined at week 0 (EDSS: 2.5; T25FW: 6.8) week 12 (EDSS: 2.5; T25FW: 6.5 [s]) and at week 24 (EDSS: 3; T25FW: 6.6). In comparison, figure 3c demonstrated a stable PwMSmod subject. This participant was examined at baseline (week 0; EDSS 3.5, T25FW: 5.4 [s]) and midway through the study^5^ (week 12; EDSS 4.5; T25FW: 4.9 [s]).

During the FLOODLIGHT study, four PwMS subjects reported relapses using the FLOODLIGHT application on their smartphone during the study. These participants’ ambulatory-based 2MWT severity estimates were investigated in figure 4.

**Figure 4:**
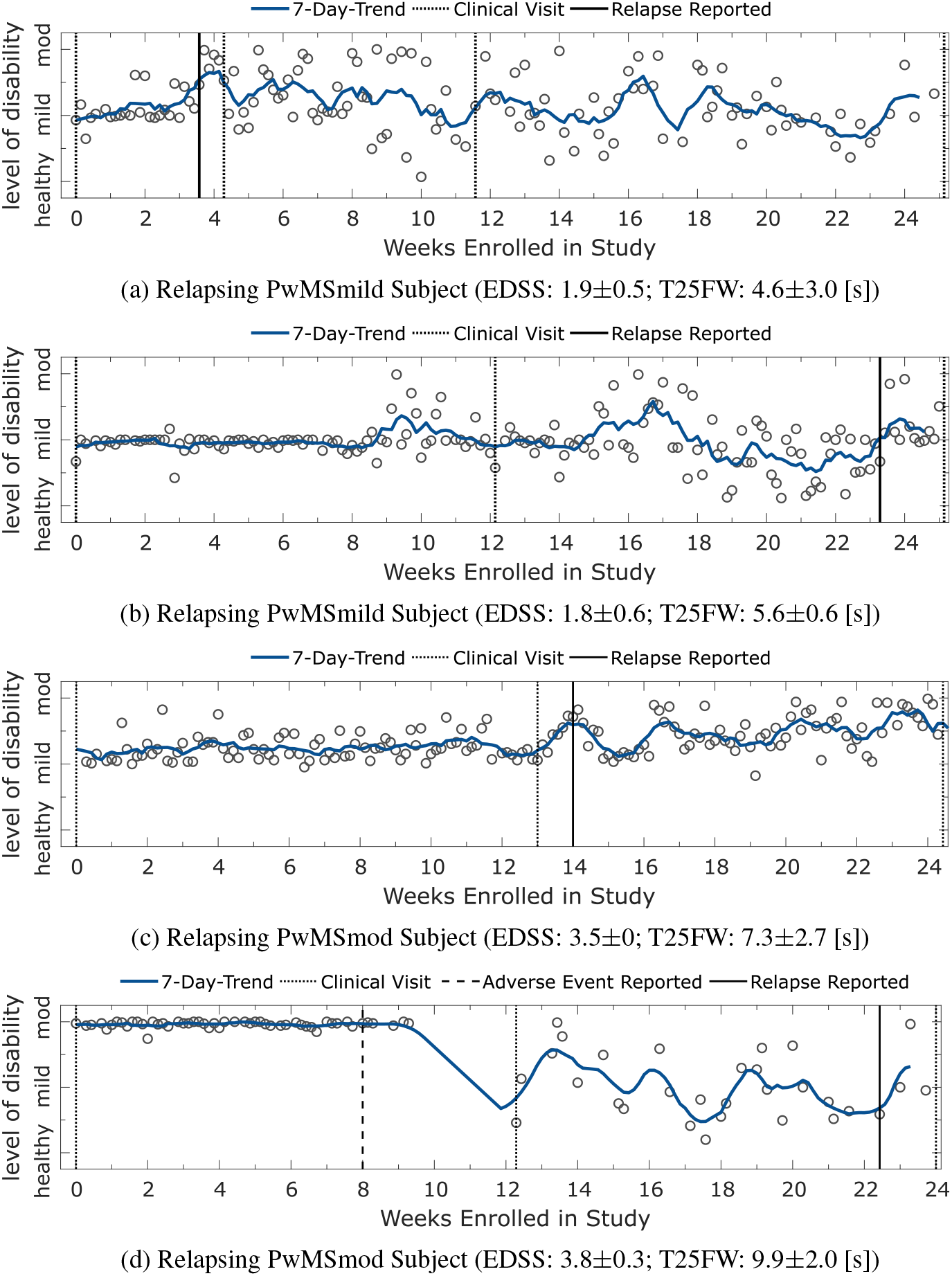
Panel plot illustrating the longitudinal severity estimate outcomes for participants who self-reported a relapse using the FLOODLIGHT smartphone application during the study. Each circle represents the severity outcome estimate for a 2MWT performed on a given date. Shaded blue lines depict the and 7-day trend, represented by the *d*-point centred moving average across days (*d*). Missing test dates (which are not depicted) were imputed using piecewise linear interpolation. Dashed black lines represent site-visits where the participant was assessed clinically. Dates of self-reported relapse onset are represented in black. Note: the participant in figure 4d also reported (non-relapse) adverse clinical events occurring on non-specified dates between weeks 8 and 12.

Figure 4a depicts the longitudinal severity outcome trend for a PwMSmild subject who reported a relapse during the FL PoC study. A black line depicts the date of relapse on-setting during week 3, which was recorded by the participant using the FLOODLIGHT application on their smartphone. Dashed black lines represent site-visits where the participant was assessed clinically. This subject was examined at baseline (week 0; EDSS 1.5, T25FW: 4.9 [s]), week 12 (EDSS 1.5; T25FW: N/A^6^) and at the study completion in week 25 (EDSS: N/A^7^; T25FW: 5.5 [s]). In week 4, 7 days after reporting a relapse, the participant was assessed during an “unscheduled visit” where they exhibited a worsening of MS symptoms, i.e. an increase in EDSS and gait related T25FW (EDSS: 2.5; T25FW: 7.5 [s]). Their relapse was evaluated as a spinal topography outbreak.

Figure 4b assesses the severity outcomes for another PwMSmild subject, with a clinical examination at baseline (week 0; EDSS: 1.5, T25FW: 4.9 [s]), and during visit 2 (week 12; EDSS: 1.5, T25FW: 6 [s]). This participant reported a relapse during week 23 where their EDSS rose by +1 during their clinical examination during study completion (EDSS: 2.5, T25FW: 5.9 [s]).

Figure 4c examines the longitudinal severity outcome trend for a PwMSmod subject who reported a relapse during the FL PoC study. A black line depicts the date of relapse on-setting during week 13, which was recorded by the participant using the FLOODLIGHT application on their smartphone. Dashed black lines represent site-visits where the participant was assessed clinically. This subject was examined at baseline (week 0; EDSS 3.5, T25FW: 4.9 [s]), midway through the study (week 12; EDSS 3.5; T25FW: 6.6 [s]) and at the study completion (week 24; EDSS: 3.5; T25FW: 10.3 [s]).

Lastly, the ambulatory severity estimates for a PwMSmod participant who self-reported a relapse is shown in figure 4d. This participant’s clinical examination was reported at baseline (week 0; EDSS: 3.5; T25FW: 7.8 [s]), mid-study as (week 12; EDSS: 4; T25FW: 10.5 [s]) and during study completion as (week 24; EDSS: 4; T25FW: 11.5 [s]). During the clinical examination in week 12, this participant also reported non-relapse adverse clinical events, occurring on unspecified dates sometime between weeks 8 and 12. As such, the time between week 8 and week 12 is marked in figure 4d beginning with a long-dashed line. This PwMSmod subject was adherent to completing daily 2MWTs, with severity outcomes estimates consistently evaluated as moderately disabled up until week 8. Thereafter, the comparative number of completed daily 2MWTs dropped dramatically until study completion. It was observed that the stability of severity estimates predicted as PwMSmod diminished, with both greater variability between severity estimates and to the adherence of the participant to complete daily 2MWTs. Furthermore, a self-reported relapse was reported by this subject during week 22 using the FL application on their assigned smartphone, as marked by the solid black line.

## 4 Discussion

The FL PoC study demonstrates the capability of smartphone-based inertial sensor measurements to monitor ambulatory-related impairments during a remotely administered 2MWT to PwMS daily over a 24 week period. In this work, it was shown how a deep network classification model could (naïvely) estimate the level of participant disability from ordinal classification categories. Severity outcome estimates stratified across HC and PwMS groups and were strongly correlated to disease status (*r* : 0.75; *ρ* : 0.71, *p <* 0.001), as measured by the EDSS – considered the ground-truth assessment in PwMS [22]. For instance, no misclassifcation of HC as PwMSmod was observed, or vice-versa, indicating that severity estimates were reflective of true disease status. More interestingly, those subjects at classification boundaries displayed severities representative of their clinical assessments. For instance, those with EDSS just above 3.5 (i.e. PwMSmod) were misclassified more as PwMSmild compared to those with EDSS much greater than 3.5, implying that a reflective estimate of disease severity could be captured by transforming a DCNN model into a simple probabilistic outcome per subject.

### 4.1 Examining Participant-level Longitudinal Trends

The longitudinal patterns of healthy controls versus participants with varying manifestations of MS severity could be characterised by examining severity outcomes over the duration of the FL study for individual subjects. For instance, figure 3 depicted examples of stable trends for correctly classified HC, PwMSmild and PwMSmod subjects respectively. While participants had some incorrect predictions, the mean severity prediction over all repeated tests reflected the participant’s true class grouping.

Evaluating subject’s performance longitudinally suggested that severity estimates may be sensitive to capture MS-symptom worsening. An intriguing observation related to the stable PwMSmod participant depicted in figure 3c, who was mainly predicted with a severity of PwMSmod, with a relatively consistent 7-day average. Some sequences of tests were predicted as milder however, particularly before the midway clinical visit in week 13. Interestingly, after week 12, this subjects’ EDSS rose by +1 to 4.5. A Brown–Forsythe (BF) test demonstrated that this subject had greater variance in their severity outcome before this clinical visit compared to after (BF, *p <* 0.01). Median severity outcomes were not significantly different between these time-points (Mann-Whitey U test, p=0.34), however mean severity outcomes were found to be significantly lower before this clinical visit than after (Welch’s *t*-test: p*<*0.05). It should be noted however that a change in EDSS scores between clinical visits did not correspond to significant changes in ambulatory-based severity estimates for all participants.

### 4.2 Examining Participant-level Relapse Events

During the FL study, four participants experienced relapses which they self-reported using the application on their smartphones. Longitudinal analysis of the trajectories of daily severity estimates from these subjects revealed useful insights into the manifestation of relapses expressed in remote inertial sensor data. For instance, two subjects displayed an increased severity outcome up to and around the data of reporting a relapse (figure 4a and 4c), suggesting that sensor-based ambulatory outcomes could potentially be sensitive enough to remotely capture relapse events.

Observing the PwMSmild participant who reported a relapse (figure 4a), severity estimates increased after reporting a relapse, which corroborated with a worsening of clinically assessed symptoms from baseline (week 0; EDSS 1.5, T25FW: 4.9 [s]) to the unscheduled clinical visit, which was prompted by the relapse (EDSS: 2.5; T25FW, 7.5 [s]). Examination of severity outcomes leading up to week 3 demonstrated consistent “mild” trends using 7-day moving averages. Interestingly, after the date of onset of self-reported relapse, severity estimates rose towards “moderate”, indicating that MS symptom manifestation had worsened. Longer term analysis demonstrated that there was a significantly higher variability in predicted severity outcomes after relapse date than before (BF, p*<* 0.001). This subject was further assessed during week 12, where their EDSS returned to as it was reported at baseline (EDSS 1.5; T25FW: N/A). Severity outcomes also returned to consistently “mild” towards the end of the study from weeks 18 onwards, where median (U test, p= 0.24) and mean (Welch’s *t*-test, p= 0.13) severity outcomes where not significantly different before- and after-relapse. This subject was predicted as PwMSmild over their entire 2MWT outcome measures.

In contrast, the example participant presented in figure 4b did not exhibit any significant changes in severity estimates around the date of reporting a relapse in week 23. However, it could also be noted that this subject’s EDSS scores rose by +1 between week 12 and 24, and their ambulatory estimated outcomes were more variable after week 12 (BF, p*<* 0.01).

Figure 4c depicted a relapsing PwMSmod subject, with severity estimates that were consistently evaluated as “mild”, up until week 13, where this participant reported a relapse on-setting using the FL application on their smartphone. Severity outcomes then increased towards “moderate” during week 13 and peaked at week 14, around the suspected relapse date reported at the end of week 13. Thereafter, severity outcomes stabilised to “mild” before becoming more variable and “moderate” until the end of the study. Considering the relapse reporting date as a threshold, it was found that severity outcomes were significantly “milder” before relapse (where severity outcomes evaluated as PwMSmild) than after relapse on-setting (where severity outcomes evaluated as PwMSmod) when testing between mean (Welch’s *t*-test, p*<* 0.001) and between median (U test, p*<* 0.001) severity outcomes. A BF test also signified that severity outcome variability was higher after relapse on-setting than before (p*<* 0.01). This subject was misclassified as PwMSmild using all available 2MWTs, but interestingly was narrowly labelled a PwMSmod and not a PwMSmild subject using their available EDSS scores (EDSS, 3.5 ± 0).

Finally, figure 4d describes the longitudinal severity outcomes for a PwMSmod participant who was consistently estimated as having moderate disability for the first 9 weeks of the study period. During the mid-way assessment at week 12, this participant recalled that non-MS related adverse clinical events had occurred at unspecified points in the previous four weeks. Interestingly, adherence to executing daily 2MWTs dropped during this period, where a long-dashed line marks the beginning between weeks 8 and 12 in figure 4d. It was observed that after week 12, the variability in sensor-based ambulatory severity estimates increased, where predictions fluctuated between healthy and moderate. Furthermore, this participant was non-adherent at providing daily 2MWTs after week 12, in comparison than the first 9 weeks. Towards the end of the study, this participant then self-reported an MS-related relapse as having occurred in week 22. As such, we need to consider not only that sensor-based outcomes could remotely evaluate a patient’s level of disability, but that an absence of available data itself might also be indicative of changes in disability status.

### 4.3 Limitations

Despite the potential of smartphone-based outcomes to remotely monitor individual participant’s ambulatory function longitudinally, there are several limitations of this study which must be considered. Importantly, the severity outcomes explored in this work were naïve estimates; although outcomes captured a trend of increased impairment with higher severity (as modelled by EDSS, figure 2), they should not be considered an exact measure of MS, nor a surrogate clinical outcome to permit any clinical diagnosis, or replace in-clinic assessments.

It should also be noted that the estimated level of participant disability were not always accurate, there were many subject misclassifications, as evident in figure 2. Particularly, some HC were incorrectly estimated as MS, as well as some PwMSmod who were underestimated to have milder level of disability. In this work, we have only shown correct and stable estimate examples, however, it must be noted that some participants, both healthy or with MS, followed irregular trends or whose estimated level of disability were consistently incorrect. It also must be acknowledged that severity estimates were based solely on 2MWT performance, an assessment originally only intended to investigate ambulatory function and fatigue in PwMS through the measurement of distance travelled [43]–[45]. Many participants in the FL PoC study may not have had ambulatory-related dysfunction, or whose milder level of disease did not affect their gait during assessment, compared to the healthy control cohort. Furthermore, the blunt demarcation of mild and moderate MS based exclusively on the clinical EDSS score – which incorporates, but is not a direct measure of ambulatory function – could lead to an unreliable assignment of those “mild” versus “moderate” MS ambulatory function. For example, some participants might exhibit “moderate” symptoms that are more apparent in other functional domains, or have subtle alteration in ambulatory ability that a remote 2MWT assessment will not be sensitive to.

In this work we proposed that averaging over categorical class predictions can create a simple and naïve estimate of ambulatory severity, but there could potentially be more informative and robust methodological approaches to learning disease severity estimates [46], [47]. It should be acknowledged that our DCNN model did not truly utilise the time-series nature of repeated 2MWT measurements from the FL PoC study. Each repeated test was treated as independent, and as such, trajectories did not incorporate any temporal information across a population or within a subject (for example, whether the previous day’s *d* − 1 test could affect the outcome at *d* or *d* + 1). It would be assumed that this is critically missing temporal information which could help build more reliable and accurate longitudinal models, and should be considered as a key next step for future work. For instance, the repeated FL assessments, and therefore sensor outcomes that were extracted, could be analysed with models that exploit this aggregation of temporal information directly [48], [49]. Another limitation of averaging posterior class predictions is that we also average over uncertain or marginal predictions, often introducing a noise and variability into the unified estimate. Indeed, constructing more robust severity outcomes would not only explore more accurate modelling techniques, but should also aim to incorporate the data captured from other functional domains in FL, such as dexterity and cognition.

Nonetheless, we believe that the work presented in this study to be of important value, emphasising the potential of remote sensor outcomes to augment current in-clinic acquired patient information. The long-term remote monitoring of PwMS function could open up the space for true personalisation: the clustering of disease trajectories or similar patients, estimating the likelihood of disease progression, quantifying response to different treatments as a population or an individual, as well catching the mutable patterns of MS disease that are only visible out-of-clinic and as a function of time.

## 5 Conclusion

This work demonstrates the capability of smartphone technologies to administer daily ambulatory assessments to patients at home, and how that sensor data recorded can be transformed through state-of-the-art deep networks, to remotely monitor ambulatory-related level of disability over a 24 week period. The rapid development of frequent, objective, and sensitive digital measures of MS disability that can be administered remotely could revolutionise routine in-clinic assessments for PwMS. In the years to come, smartphone-based outcomes may identify and monitor digital signs of MS-related degeneration, ultimately informing better disease management techniques, to learn how different patients respond to various treatments, and potentially enabling the development of personalised therapeutic interventions.

## Data Availability

Qualified researchers may request access to individual patient level data through the clinical study data request platform (https://vivli.org/). Further details on Roche's criteria for eligible studies are available here (https://vivli.org/members/ourmembers/). For further details on Roche's Global Policy on the Sharing of Clinical Information and how to request access to related clinical study documents, see here (https://www.roche.com/research_and_development/who_we_are_how_we_work/clinical_trials/our_commitment_to_data_sharing.htm).

## Acknowledgements

The authors would like to thank all staff and participants involved in capturing test data. This study was sponsored by F. Hoffmann-La Roche Ltd. This research was supported by the National Institute for Health Research (NIHR) Oxford Biomedical Research Centre (BRC). This research also received funding from the Flemish Government under the “Onderzoeksprogramma Artificiële Intelligentie (AI) Vlaanderen” programme. During the completion of this work, A. P. Creagh was a Ph.D. student at the University of Oxford and acknowledges the support of F. Hoffmann-La Roche Ltd.; F. Dondelinger and F. Lipsmeier are employees of F. Hoffmann-La Roche Ltd; M. Lindemann is a consultant for F. Hoffmann-La Roche Ltd. via Inovigate; M. De Vos has nothing to disclose.

## Footnotes

3 Note: an EDSS of zero in this case refers to a normal neurological exam, the subject is healthy and has no disability.

4 N/A denotes scores not assessed at this visit.

5 Note: clinical assessment scores were not made available for this participant at study completion.

6 See footnote 4

7 See footnote 4

## Notes

### Clinical Protocols

https://clinicaltrials.gov/ct2/show/NCT02952911

### Author Declarations

All participants provided informed consent, and the ethical approval was obtained from ethics committee of the Hospital Universitari Vall d'Hebron, Barcelona, Spain and the institutional review board of the University of California San Francisco, San Francisco, CA, USA, prior to study initiation. The study was registered on clinicaltrials.gov (NCT02952911).

